# Cognitive Therapy Approach For Post-Stroke Patients : A Review Of Literature

**DOI:** 10.1101/2023.12.15.23300013

**Authors:** Sarida Surya Manurung, Moses Glorino Rumambo Pandin

## Abstract

**Introduction:** Stroke is a highly consequential medical condition, characterized by a substantial risk of death and disability. Based on the data of the World Health Organization (WHO), stroke was contibuted for 6.2 million of mortalities, the second cause of death globally. In particular, half of stroke survivors encounter challenges in performing daily activities, and the psychosocial aspects of their experience often lead to a diminished quality of life, contributing to conditions like depression. In addition for the impact of physical, stroke also can induce the cognitive barriers, impacted to an attention, orientation, retention, and cognitives functions. In addressing these cognitive challenges, particularly through cognitive therapy, was shown promising in reducing levels of anxiety and depression among post-stroke individuals. This literature research deals to examine research outcomes related to various post-stroke cognitive therapies. The objective is to describe the advantages and disadvantages of this therapeutic approaches, clarify the effectiveness in rehabilitate of cognitive and psychological consequences of stroke.

**Method:** Literature obtained through electronic media on Science Direct, Scopus and Google Scholar by used the keyword Cognitive Therapy Approach in Post Stroke Patients and it was found 5 articles that met with the criteria which had been published less than the last 10 years.

**Results:** this study shows that cognitive interventions that duplicate memory, processing speed and attention can produce significant improvements in several cognitive domains. This therapy teaches compensatory strategies such as using a notebook or daily planner and analyzing tasks logically until activities are carried out well in daily life which involves exercises to increase attention and requires internal neurological attention. The function of this training includes visual and auditory skills, both of which are important for everyday training and information processing designed to improve retention and recall of information and improve memory. Attention and executive skill function are interdependent and have a significant impact on daily functioning. Therefore, exercises that improve attention, working and short-term memory can improve general mental abilities and improve a persons ability to process information.

**Conclusion:** The results of this study indicate that cognitive therapy can help the patients to achieve the recovery optimally both in cognitive or emotional aspects.

## Introduction

Stroke is a very serious disease due to high mortality and disability (Burton et al., 2013). In 2017, WHO stated that stroke was the second cause of death in the world, resulting in 6.2 million people dying. Overall, 30% of the 60% of survivors worldwide suffer from post-stroke (PSD) within 1 year after stroke. After a stroke, 35% of patients experience functional dependence, which indicates that the stroke is the main cause of disability and the other half have difficulty carrying out daily routines. The psychosocial problems of stroke sufferers have a negative impact on quality of life. Emotional adjustment is associated with changes in coping strategies related to quality of life. In addition, it has been proven that caregivers also experience a large burden in their partner’s emotional problems. Cognitive and emotional complaints are often found in patients in the chronic phase after stroke, if this does not happen the symptoms become chronic (Rinjani, S.at al.2021).

Post-stroke patients often experience cognitive decline as a result of a stroke that occurs in the brain’s blood vessels and has a fairly high incidence rate in adults and the elderly (Levine et al., 2015). Cognitive obstacles after stroke are related to long-term survival which leads to the patient’s quality of life (Health-related Quality of Life), which is the result of cognitive disorders and other major functions that can interfere with daily life activities and often cause the sufferer to become dependent on other people. others, as well as reducing work productivity which consists of 6 items related to cognitive evaluation for stroke sufferers (Rahayu et al., 2014).

For post-stroke patients, medical rehabilitation interventions are needed so that they are able to independently care for themselves and carry out daily life activities without having to continue to be a burden on their families. However, not all patients have the opportunity to continue the stroke rehabilitation program after returning home from treatment. Most of this is due to the unavailability of medical rehabilitation facilities near where the patient lives. In general, subacute and chronic phase stroke rehabilitation can be handled through simple medical rehabilitation procedures that do not require sophisticated equipment. Focusing on efforts to prevent immobilization complications which can have an impact on worsening conditions and restoring independence in daily activities, it is hoped that patients can achieve a better quality of life. Primary Health Care plays a very important role (Wirawan, 2009).

To avoid this problem, cognitive behavioral therapy should be a solution. Cognitive therapy is individual psychotherapy carried out to train clients to change the way clients explain and see things when they experience disappointment, so that clients feel good and act more productively (Townsend, 2005). Through cognitive therapy, individuals are taught and trained to control distorted thoughts or ideas properly and consider developmental factors and mood disorders. Research on cognitive therapy has been carried out by Rahayuningsih, Hamid, Mulyono (2007); Kristyaningsih, Keliat and Helena (2009) and cognitive applications have been carried out by Sartika, Hamid and Wardani (2011), Jumaini, Hamid and Wardani (2011); Syarniah, Hamid and Susanti (2011) showed the results that cognitive therapy has an effect on changes in a person’s selfesteem and cognitive independence.

## METHODE

The literature observation method was used to gain a deeper understanding by searching the literature related to the research theme. Literature research were carried out through electronic sources such as Scopus, Google Scholar, EBSCOhost, PubMed, and Science Direct. The keywords used in the literature research were “Cognitive Therapy Approach for Post-Stroke Patients”. In the first search with that keywords were found 110 articles related to the problem. Next, the further research was carried out by limiting the range of years publication between 2013 and 2023, till 65 articles were met to this criteria. The aim of limitation of years’ publication is to keep writing up-to-date based on the latest research results. After reading and selecting, 5 articles were found that were in accordance with the research theme. The selection of articles is based on relevance criteria, including clarity of source, originality of the article, suitability to the research topic, publication year within the last 10 years, and the presence of main content regarding Cognitive Therapy, post-stroke, and Cognitive Therapy Approaches.

## RESEARCH RESULT

The author was found 110 articles from databases, including Scopus: 15 articles; Published: 35 articles; google schoolar: 15 articles, After removing duplicates and applying inclusion and exclusion criteria, the final sample consisted of 5 studies. Search and self-selection processes in the general outline of learning (Figure 1).

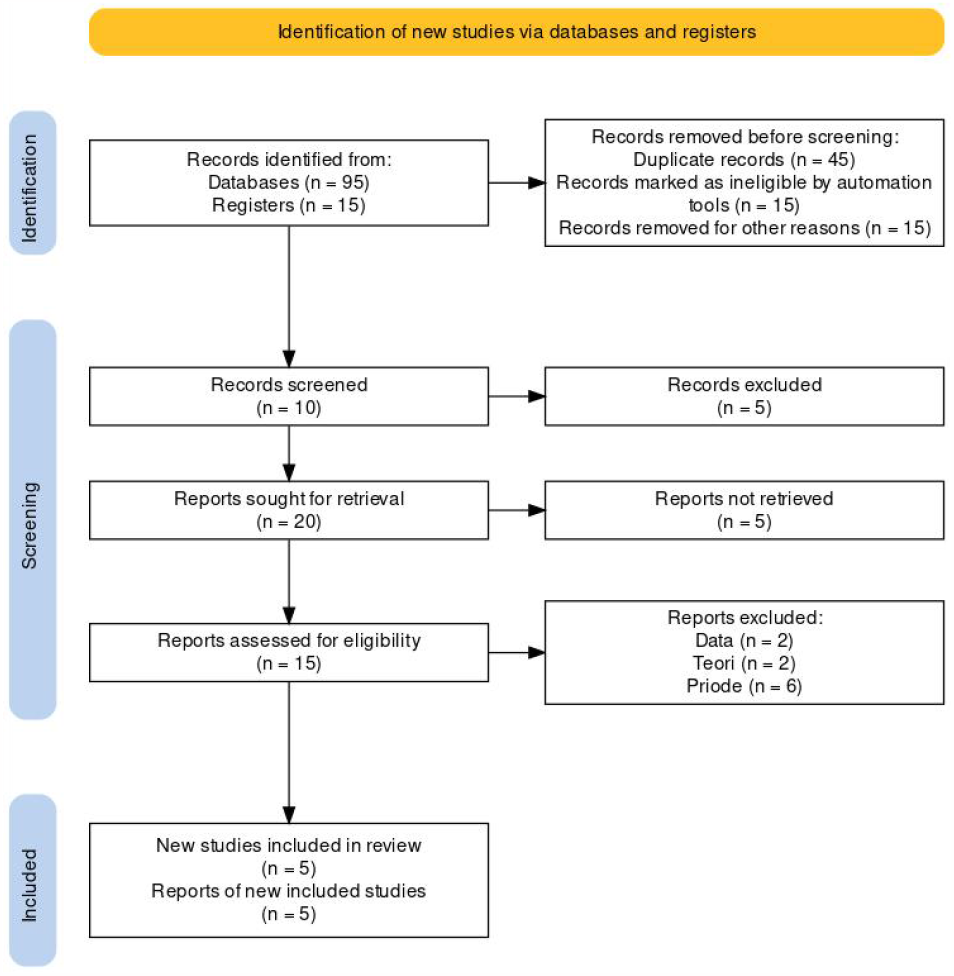

This research was carried out by using the literature review method, namely gaining more understanding by looking the literature that related to the research theme. A literature search was carried out on electronic media (internet) on the electronic databases Google Scholar, ebscohost, PubMed and Science Direct. The keywords for searching the literature were “cognitive behavior therapy in patients with stroke and depression.” in he first searching by using that keywords, it was found 110 articles that related to the theme. Then, the further search was carried out by limiting the range of years publication between 2013 and 2023, till 65 articles were met to this criteria.

The carried out of years’ articles restriction was to maintain the writing still up -to -date based on the latest research results. After reading the articles and selecting them according to the research theme, it was found 5 articles that suitable with the theme. The reason for the relevance of the selected articles because the source or the article was clear, original, related to the desired topic, the publication year was less than 10 years and the article had the main content, namely post-stroke, stroke, cognitive therapy and Cognitive Therapy In Post Stroke

**Table 1.**
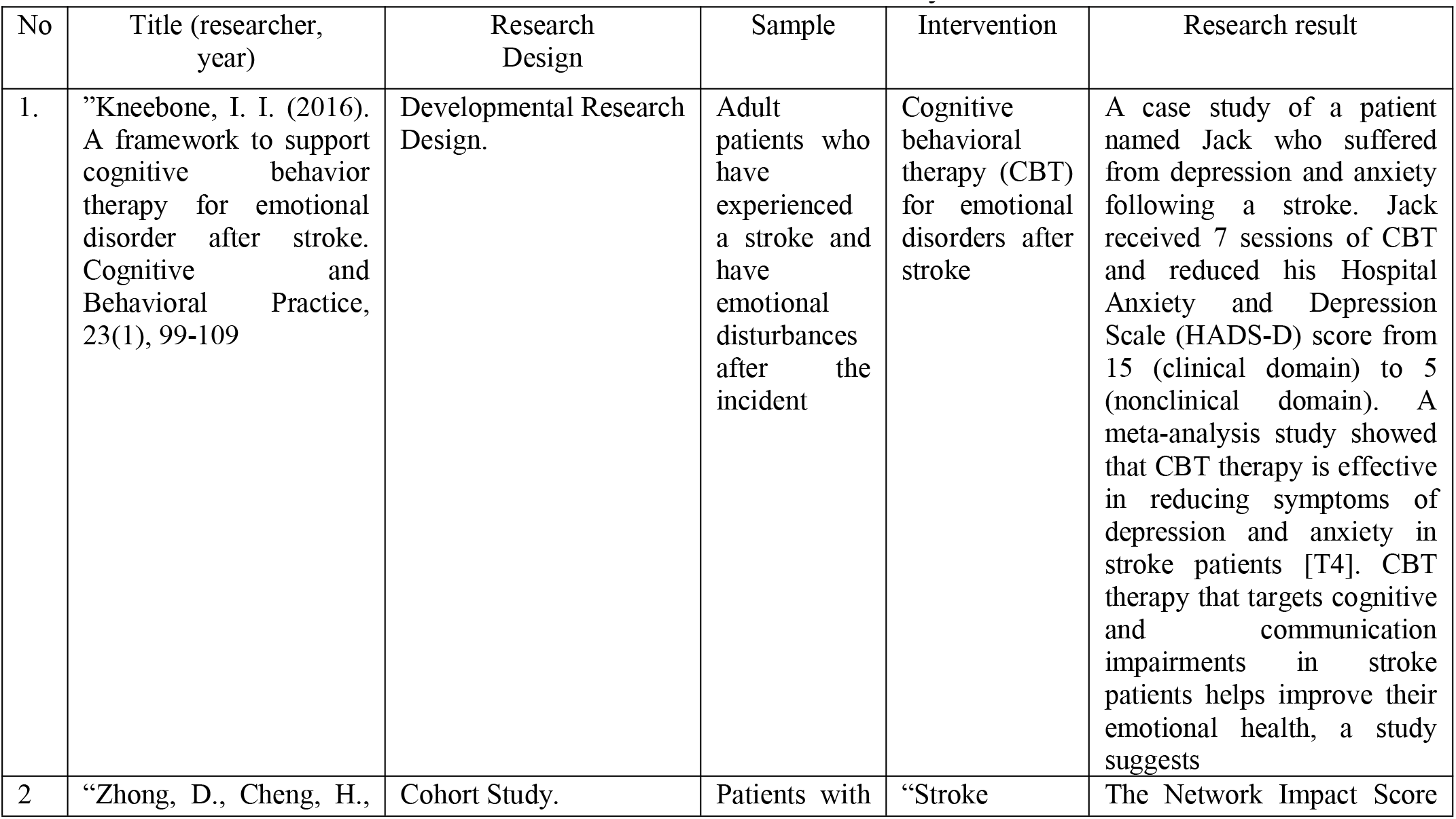
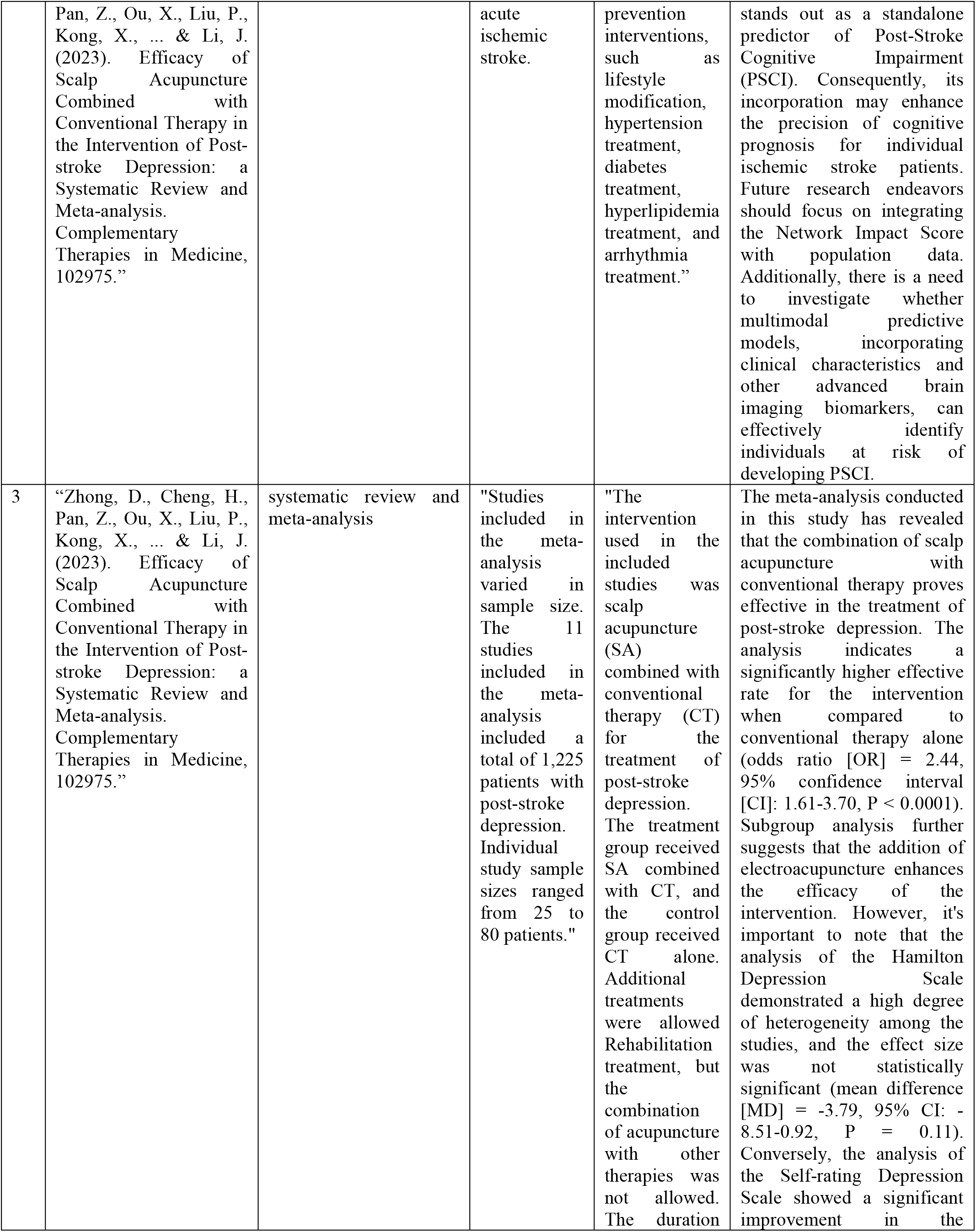
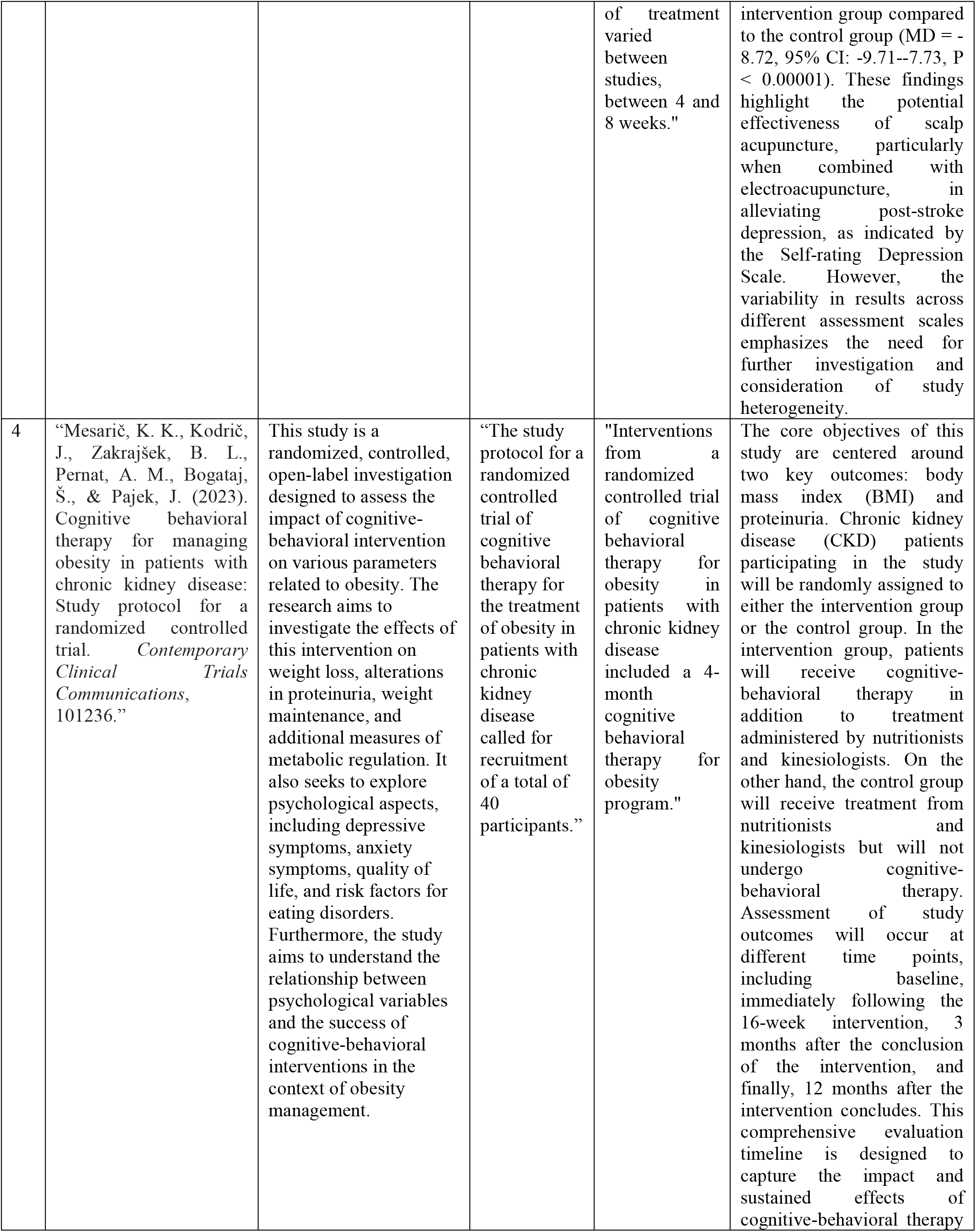
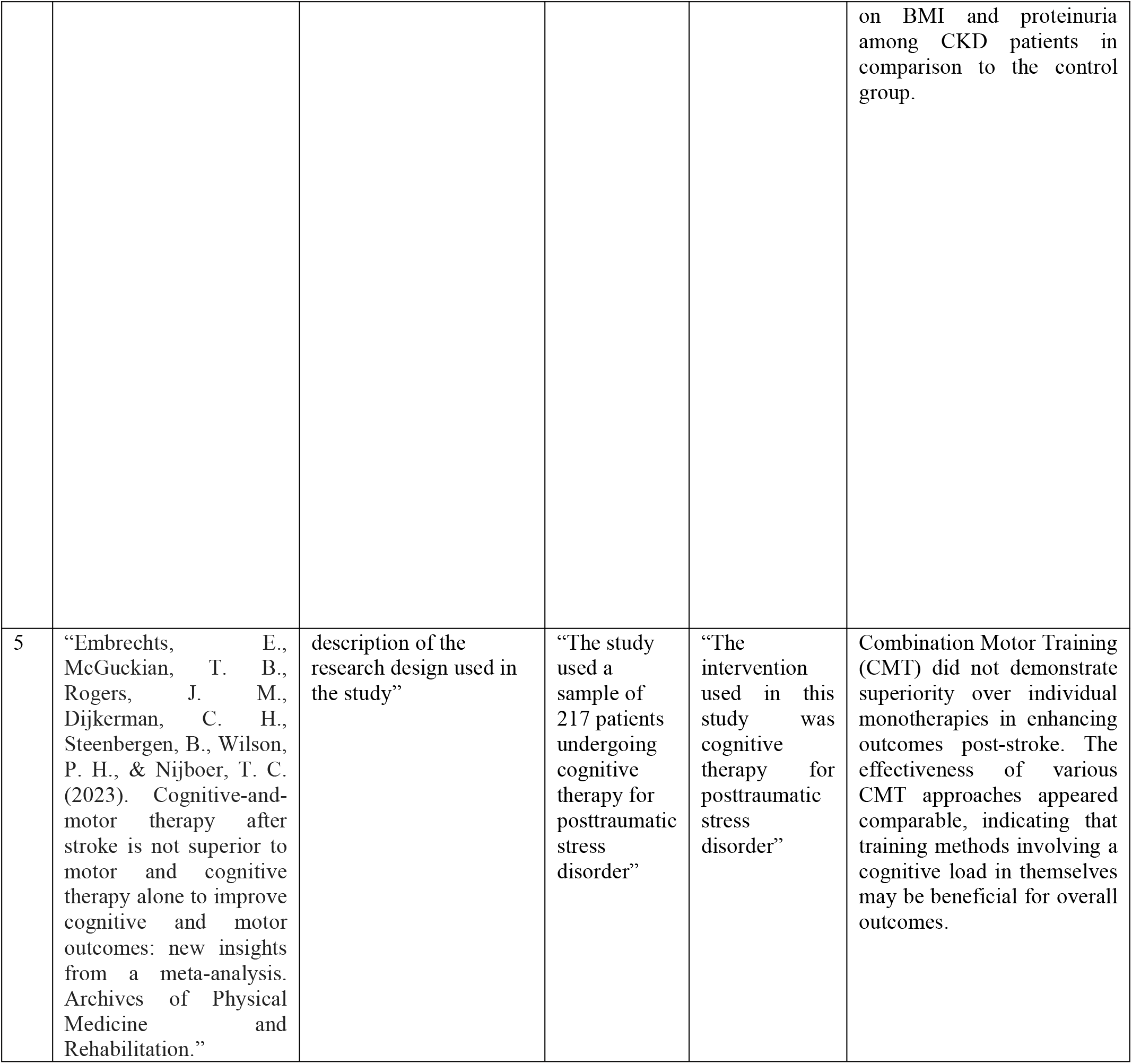
Characteristics Study.

## DISCUSSION

In summary, the collective findings that the articles were presented above suggest that the Cognitive Therapy Approach for Post-Stroke Patients is widely regarded as highly effective. This therapeutic approach is designed to address and mitigate cognitive challenges that may manifest following a stroke. Considering that strokes can effected to the brain damage, has crucial impact to cognitive functions like memory, attention, problem-solving, and abstract thinking, the application of cognitive therapy emerges as a valuable intervention method for managing these post-stroke cognitive issues. Based on Kneebone’s research, I.I. (2016). The effectiveness of Cognitive Behavioral Therapy (CBT) for individuals dealing with emotional issues after a stroke extends beyond its impact on emotional outcomes. The positive effects of CBT are not confined solely to emotional well-being; rather, they also contribute significantly to functional recovery. This implies that the application of CBT not only addresses and improves emotional states in post-stroke patients but also plays a crucial role in enhancing their overall functional abilities and recovery. This is in line with the research results of Biesbroek, J.M., Weaver, N.A., Aben, H.P., Kuijf, H.J., Abrigo, J., Bae, H.J.,.&Biessels, G.J. (2022). Cognitive Behavioral Therapy (CBT) has demonstrated efficacy in alleviating symptoms of depression and anxiety. This therapeutic approach is frequently employed as the primary method for treating depression due to its effectiveness. By focusing on modifying negative thought patterns and behaviors, CBT aims to reduce depressive symptoms and alleviate anxiety. As a result, CBT stands out as a widely utilized and successful form of therapy for individuals dealing with depression, offering a practical means of addressing and managing both depressive and anxiety-related symptoms. (Forman, Herbert, Moitra, Yeomans, & Geller, 2007). The meta-analysis in the study revealed that the combination of scalp acupuncture and conventional therapy proved to be an effective approach in addressing post-stroke depression. The findings suggest that when scalp acupuncture is integrated with traditional therapeutic methods, it demonstrates effectiveness in the treatment of depression following a stroke. This combination approach appears to offer a valuable and promising strategy for managing post-stroke depression.. Zhong, D.at al. (2023). This trial is structured as a randomized, controlled, open-label study with the primary objective of examining the effects of a cognitive-behavioral intervention on various dimensions of obesity management. The study encompasses an assessment of outcomes such as weight loss, alterations in proteinuria, weight maintenance, and diverse metabolic and psychological parameters. Additionally, the research endeavors to gauge the impact of the intervention on secondary factors, including depressive symptoms, anxiety levels, quality of life, risk factors for eating disorders, and the potential correlation between psychological variables and the success of cognitive-behavioral interventions. Through this comprehensive approach, the study seeks to provide of understanding the multifaceted effects of cognitive-behavioral interventions in the context of obesity management thoroughly. Cognitive-behavioral therapy (CBT), a form of psychotherapy, is a central component of the intervention. CBT focuses on cognitive processes, beliefs, assumptions, and behaviors with the goal of influencing and improving emotional wellbeing. By targeting negative, irrational, and disruptive belief systems, CBT aims to facilitate a gradual shift toward healthier and more normal social reactions and behaviors. The provision of cognitive-behavioral therapy in this context is anticipated to offer poststroke patients a more effective path to recovery and improved overall well-being. (Lincoln &Flannaghan, 2010). Changes in a person’s cognitive aspects will be followed by their behavior (Susana, Parmadi, &Adi, 2015). This statement is appropriated with Beck (2011), that negative thoughts will increase a person’s tendency to experience depression when experiencing a stressful event, so that they can change their way of thinking to be more adaptive, which is one way to reduce it. level of depression (Beck, 2011; Fitriani, 2018).

## Conclusion

Based on several presentations of research results related to the Cognitive Therapy Approach for Post-Stroke Patients, it can be concluded that behavioral cognitive therapy is able to reduce anxiety for post-stroke patients significantly by seeing a decrease in anxiety levels. Besides that, by observing and identifying changes in aspects of the subject’s thoughts, feelings, physiology and behavior, family support is needed to change the negative thoughts of post-stroke patients to more positive ones. So that they can increase their motivation and expectation for recovery by carrying out cognitive behavioral therapy that collaborate with health workers to provide the education.

## Data Availability

All data produced in the present work are contained in the manuscript

## Notes

### Competing Interest Statement

The authors have declared no competing interest.

### Funding Statement

This study did not receive any funding

